# Oral health inequality in people with severe mental illness: a cross-sectional study using National Health and Nutrition Examination Survey 1999-2016

**DOI:** 10.1101/2021.03.17.21253840

**Authors:** Jing Kang, Jianhua Wu, Vishal Aggarwal, David Shiers, Tim Doran, Jasper Palmier-Claus

## Abstract

**OBJECTIVE:** To explore whether people with severe mental illness (SMI) experience worse oral health compared to the general population, and the risk factors for poor oral health in people with SMI.

**METHOD:** This study used cross-sectional data from the National Health and Nutrition Examination Survey (1999-2016) including on self-rated oral health, ache in mouth, tooth loss, periodontitis stage, and number of decayed, missing, and filled teeth. Candidate risk factors for poor oral health included demographic characteristics, lifestyle factors, physical health comorbidities, and dental hygiene behaviours. The authors used ordinal logistic regression and zero-inflated negative binomial models to explore predictors of oral health outcomes.

**RESULTS:** 53,348 cases were included in the analysis, including 718 people with SMI. In the fully adjusted model, people with SMI were more likely to suffer from tooth loss (OR 1.40, 95% CI: 1.12-1.75). In people with SMI, the risk factors identified for poor oral health outcomes were older age, white ethnicity, lower income, smoking history, and diabetes. Engaging in physical activity and daily use of dental floss were associated with better oral health outcomes.

**CONCLUSIONS:** People with SMI experience higher rates of tooth loss than the general population, and certain subgroups are particularly at risk. Having a healthy lifestyle such as performing regular physical exercise and flossing may lower the risk of poor oral health. These findings suggest opportunities for targeted prevention and early intervention strategies to mitigate adverse oral health outcomes.

**Significant outcomes (x3):**

1. People with severe mental illness were at 40% higher risk of tooth loss when compared to the general population.
2. Older adults, smokers and people with diabetes were at particularly high risk of poor oral health.
3. Physical exercise and daily use of dental floss were associated with better oral health outcomes.

**Limitations (x3):**

1. The number of cases with data on periodontal disease was limited.
2. The study was cross-sectional so causation could not be inferred.
3. The analysis used prescriptions of antipsychotic and mood stabilising medication as a proxy measure of severe mental illness, as clinical diagnoses were not available in the dataset.

**Data availability statement:** The NHANES 1999-2016 data is available at CDC website: https://www.cdc.gov/nchs/nhanes/index.htm, and is accessible and free to download for everyone.

## Introduction

Poor physical health in people with severe mental illness (SMI; e.g. psychosis, bipolar disorder) is a major research priority (1). One important, but often neglected, area of investigation is the disparity in oral health (2). Initial research suggests that people with SMI may experience worse oral health outcomes compared to the general population (3) with higher rates of decayed, missing, and filled teeth (4). In some cases, poor oral health can interfere with basic functions, such as eating (5). There is evidence in the general population that poor oral health can have profound impact of quality of life (6) and can limit employment opportunities (7, 8). There is a major need for a greater understanding of oral health inequalities in SMI in order to develop more effective and targeted interventions.

We do not yet understand the reasons for poor oral health in people with SMI. There is some indication that patients are less likely to brush their teeth or own a toothbrush compared to the general population (9-11), and that they have elevated risk factors including smoking (12), drug use (13), and poor diet (14). Xerostomia is a common side effect of psychotropic medication and can elevate risk of tooth decay and infection (15). Access to treatment may also be limited in SMI, with one study in Denmark suggesting that only a third of patients will attend an annual dental appointment (16). There is evidence that both cardiovascular disease (CVD) and diabetes are associated with poor oral health (17, 18). Not only are these health conditions highly prevalent in people with SMI (19, 20), but when combined with poor oral health they also add to the burden that many may face in managing multiple morbidities, treatments and healthcare providers.

To date, research on oral health outcomes in SMI have mostly used small unrepresentative samples without controlling for relevant clinical and demographic covariates. This prevents us from accurately knowing the size of oral health inequalities in SMI, compared the general population. Additionally, little is known about the factors contributing to poor oral health in SMI. In this study, we explore oral health inequalities in a large scale, representative sample, the National Health and Nutrition Examination Survey (NHANES) (21, 22).

### Aims of the study

The aims of this study are to explore whether people with SMI have worse oral health (dentition, dental caries, periodontal status, and self-reported oral health status) compared to people without SMI. We also sought to identify risk factors including demographics, lifestyles, comorbidities and oral hygiene behaviours, for poor oral health outcomes in people with SMI. Taken together, we intended to understand the size and causes of poor oral health in SMI.

## Methods

### Study design

The study followed the STROBE guidelines. Study participants came from cross sectional NHANES 1999-2016. NHANES is a national survey designed to assess the health and nutritional status for the non-institutionalised United States population using a stratified, multistage, probability sampling design. NHANES has been using the same survey structure and conducting data collection in two-year cycle since 1999/2000, and consists of extensive anthropometric, socioeconomic, health and dental related examinations and questionnaires, as well as laboratory testing for biomarkers. Height, weight and waist circumference were measured onsite by trained examiners; dental-related measures were taken by trained dental survey staff and periodic quality controlled by a second “gold standard” examiner. The methods and design for the survey are available elsewhere (21).

### Study participants

We extracted data from nine NHANES surveys between 1999 and 2016. The resulting sample from NHANES for participants over 18 years old was 53,348 participants (25,709 men and 27,639 women). NHANES does not contain clinical diagnoses of mental illness, so participants’ prescription medicine in the past month were extracted and people with SMI were identified based on the type of medication that they were taking. If participants reported to take one or more of the medications presented in Appendix I, we considered them have severe mental illness. Validation of drug code is via ICD-10-CM code, and mental illness such as bipolar, psychosis and schizophrenia were identified.

### Oral health outcome measures

Oral health outcomes include dentition, dental caries, periodontal status, and self-reported oral health status.

- Dentition was measured by the number of teeth as count between 0 (edentulous) and 32 (full dentition) by trained and calibrated health technologists. Tooth loss due to traumatic injuries were excluded in the analyses because we focus on oral disease in this study. For the analysis, the tooth loss status is derived from the number of teeth and categorized as ‘no loss, 1-10, 11-20, 21-31 and edentulous’.
- Dental caries is reported as number of decayed, missing, or filled teeth (DMFT) and is derived from the coronal caries status of each tooth when coded as “missing due to dental disease, permanent tooth with a restored surface condition, permanent root tip is present but no restorative replacement is present, missing due to dental disease but replaced by a removable restoration, missing due to dental disease but replaced by a fixed restoration, permanent tooth with a dental carious surface condition”. Number of coronal decayed teeth (DT) is derived from the same tooth coronal caries status when coded as “permanent tooth with a dental carious surface condition”, and number of missing teeth (MT) due to decay is obtained when the coronal caries status was coded as “missing due to dental disease, missing due to dental disease but replaced by a removable restoration, missing due to dental disease but replaced by a fixed restoration”.
- Periodontal status was classified as ‘none, mild, moderate and severe’ by using standard case definition for surveillance of periodontitis (21). Severe periodontitis was defined as having two or more sites with >=6mm clinical attachment loss and one or more sites with >=5mm pocket depth (not on the same tooth). Moderate periodontitis was defined as two or more sites with >=4mm clinical attachment loss or two or more sites with pocket depth of >=5mm (not on the same tooth). Mild periodontitis was defined as two or more sites with >=3mm clinical attachment loss and two or more sites with >=4mm pocket depth (not on the same tooth), or one site with >=5mm pocket depth.
- Self-reported oral health included self-rated oral health status and ache in mouth. Self-rated oral health status was obtained in the interview question “How would you describe the condition of your teeth and gums?”, with the options ‘Excellent’, ‘very good’, ‘good’, ‘fair’, and ‘poor’. Ache in mouth was obtained from the interview question “How often during the last year has you had painful aching anywhere in your mouth?” with options ‘very often’, ‘fairly often’, ‘occasionally’, ‘hardly ever’, and ‘never’.

### Exposures

The following set of variables were included in the analyses:

- Demographic variables: Age (18 and above, scale), sex (male or female), ethnicity (white or other race), education qualification (high school or below, college or above), marital status (not married, married), and ratio of family income to poverty (scale).
- Anthropometric measures: body mass index (BMI, kg/m^2^), waist group (low (men <=94cm, women<=80cm), high (men 94-102cm, women 80-88cm), very high (men >102cm, women >88cm).
- Lifestyle factors: smoking status (non-smoker, ex-smoker, current smoker), cigarette number in the past 30 days (scale), had at least 12 alcohol drinks in one year (yes or no), substance misuse (ever used cocaine or other street drug, yes or no), moderate physical activity over past 30 days (yes or no); sugar intake (gram), carbohydrate intake (gram), and energy intake (KCAL).
- Comorbidities (yes or no): cardiovascular disease (including with at least one of congestive heart failure, coronary heart disease, angina, heart attack, and stroke) and diabetes.
- Dental hygiene behaviour: Time since last dental visit (less than 1 year, over 1 year, never), tooth brush frequency per day (once or less, twice or more), use dental floss (no, not everyday, everyday).

### Statistical analyses

Descriptive statistics were performed to compare people with and without SMI, concerning demographics, anthropometrics, lifestyles, comorbidities, dental hygiene behaviour, and all oral health outcomes. Continuous variables were presented as mean (SD) or median (interquartile range) and categorical variables were reported as frequency (%). The general population were matched to people with SMI on a ratio 1 to 3 based on their age and gender because the distribution for people with and without SMI were different in the original dataset (people with SMI were of older age compared with people without SMI), and matching the sample would provide comparable results.

For statistical modelling, self-rated oral health was further grouped as ‘excellent or very good or good’ and ‘fair or poor’, ache in mouth as ‘never or hardly ever’ and ‘occasionally to very often’, and periodontal status was further grouped as ‘none’, ‘mild to moderate’, and ‘severe’ to balance the proportions in each category. Smoking was grouped as two categories (non-smoker, ever smoker) as was dental visiting (less than 1 year, more than 1 year or never). Alcohol and energy intake were excluded in the statistical model because they were not significant between people with and without SMI. Education was highly correlated with family income and therefore only the latter was included in the model. Cigarette number only applied to smokers so it was excluded in the modelling process. Tooth brushing frequency had only a very small number of responses (<1% of the total sample) in both groups so it was excluded. The outcome variables of dentition (tooth loss) and periodontal status were ordinal variables necessitating ordinal regression models. Self-reported oral health (self-rated oral health status, ache in mouth) were binary so logistic regression models were applied. First, a univariable model with one of the ordinal oral health outcomes as dependent variable and group (SMI or non-SMI) as independent variable was performed. Then multivariable models were performed with gradual adjustment of demographic, lifestyles, comorbidities, and dental hygiene behaviours. Similarly, zero-inflated negative binomial (ZINB) models were used to compare SMI and non-SMI population on dental caries (DMFT, DT and MT) experience, because dental caries variables have excess zeros and follows a negative binomial distribution. (Figure 2) ZINB model is a 2-part model, with logit model predicting excessive zeros, and negative binomial model predicting the counts (23). Similar approaches for the univariable ZINB model and multivariable ZINB model were applied for DMFT, DT, and MT, respectively.

**Figure 1.**
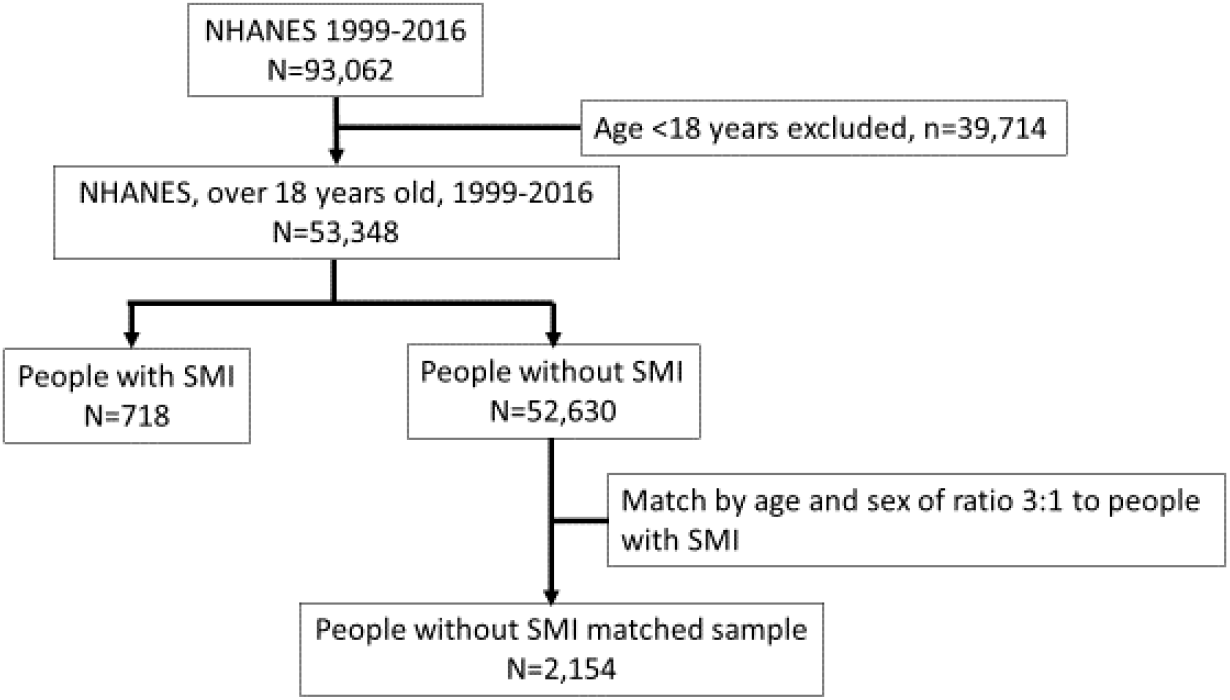
Flowchart of the participants selection.

**Figure 2.**
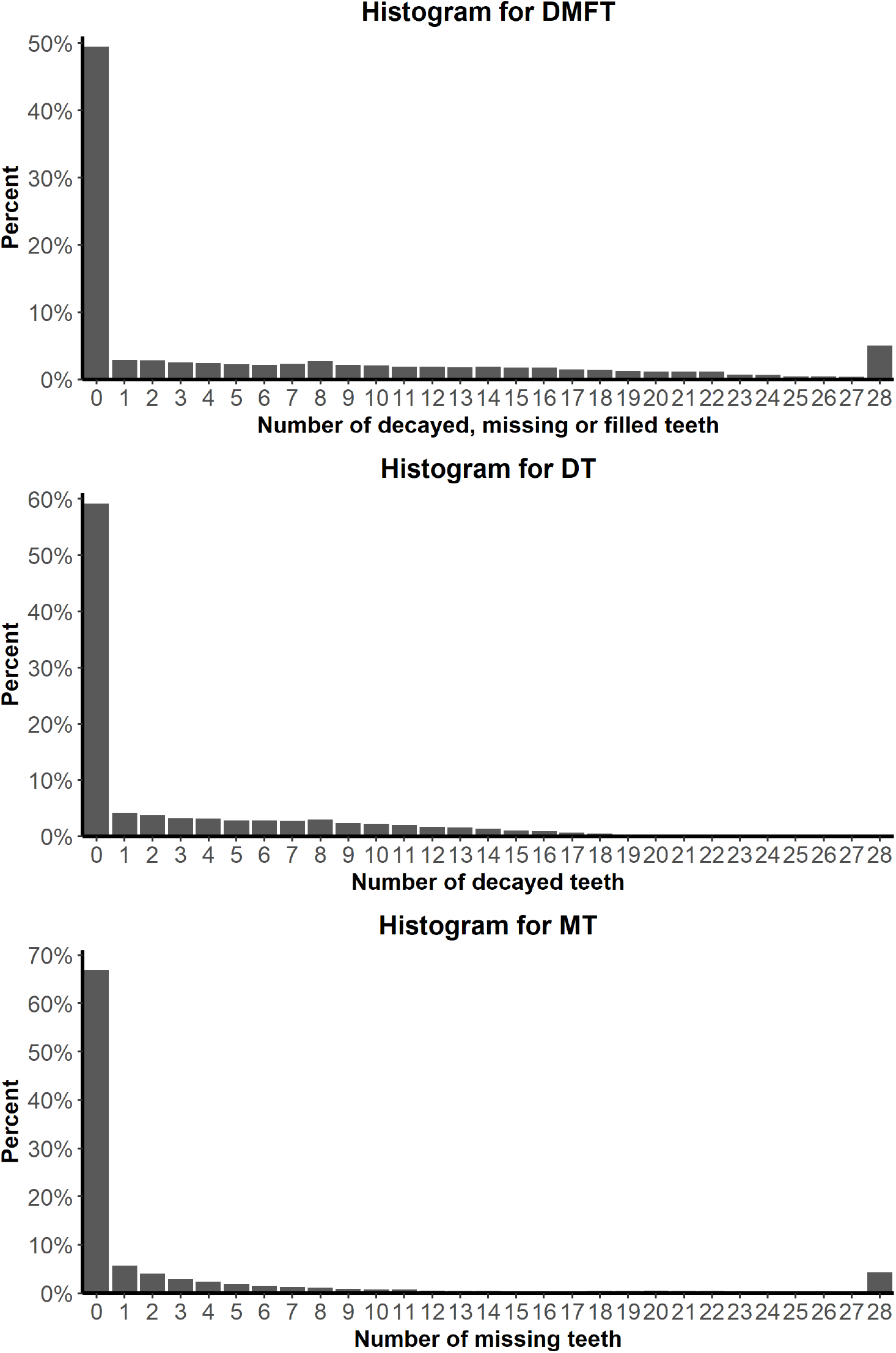
Distributions of the number of decayed, filled and missing teeth (DMFT), the number of decayed teeth (DT), and the number of missing teeth (MT).

When investigating predictors of oral health status in people with SMI alone, ordinal regression, logistic regression, and ZINB models were used for ordinal, binary, and scale oral outcomes respectively with similar approach. Missing data were imputed five times through multiple imputation by chained equations. Pooled modelling estimates and accompanying standard errors (SE) were generated according to Rubin’s rules.(24). Statistical analyses were performed in R version 3.4.1 (https://cran.r-project.org/) with various packages.

## Results

53,348 participants were included in the analyses. Figure 1 shows how participants were selected from NHANES. The average age of the total sample was 47.5 years (SD 19.6 years) and 25,709 (48.2%) were men.

### Characteristics between people with and without SMI

The mean age of people with SMI was 51.1 (*SD* 16.8) years, higher than general population sample (mean age: 47.4 years, *SD* 19.6, *p*<0.01). A higher proportion of people with SMI were white (52.2% *vs* 43.7%, *p*<0.001), and less people with SMI had higher education degree (41.3% vs 48.6%, *p*<0.001). More people with SMI were unmarried (70.6% *vs* 49.3%, *p*<0.001) and their family income was lower (ratio to poverty 1.8 vs 2.5, *p*<0.001; Table 1)

**Table 1.** Characteristics of participants with and without SMIs. NHANES (n = 53,348), 1999-2016.

| *Characteristic* | *Total* | *Without SMIs* | *With SMIs* | *p-value* |
| --- | --- | --- | --- | --- |
| *n* | 53348 | 52630 | 718 |  |
| *Age in years, mean (SD)* | 47.5 (19.6) | 47.4 (19.6) | 51.1 (16.8) | <0.001 |
| *men* | 25709 (48.2) | 25378 (48.2) | 331 (46.1) | 0.275 |
| *Ethnicity, white* | 23388 (43.8) | 23013 (43.7) | 375 (52.2) | <0.001 |
| *Education, college or above* | 23974 (48.5) | 23691 (48.6) | 283 (41.3) | <0.001 |
| *Marital status, not married* | 25418 (49.6) | 24923 (49.3) | 495 (70.6) | <0.001 |
| *Ratio of family income to poverty, mean (SD)* | 2.5 (1.6) | 2.5 (1.6) | 1.8 (1.4) | <0.001 |
| *BMI (kg/m<sup>2</sup>), mean (SD)* | 28.6 (6.8) | 28.6 (6.7) | 30.5 (7.6) | <0.001 |
| *Waist circumference <sup>a</sup>* |  |  |  | <0.001 |
| *Low* | 13215 (27.5) | 13112 (27.7) | 103 (17.3) |  |
| *High* | 9291 (19.4) | 9204 (19.4) | 87 (14.6) |  |
| *Very high* | 25473 (53.1) | 25066 (52.9) | 407 (68.2) |  |
| *Had at least 12 alcohol drink in one year* | 30261 (69.4) | 29865 (69.4) | 396 (67.9) | 0.454 |
| **Smoking status** |  |  |  | <0.001 |
| **Non-smoker** | 30575 (57.3) | 30305 (57.6) | 270 (37.6) |  |
| **Ex-smoker** | 12278 (23.0) | 12105 (23.0) | 173 (24.1) |  |
| **Current smoker** | 10495 (19.7) | 10220 (19.4) | 275 (38.3) |  |
| **Cig number in past 30 days, mean (SD)** | 12.6 (10.6) | 12.5 (10.5) | 16.4 (12.8) | <0.001 |
| **Substance Misuse (cocain/herion)** | 5766 (17.6) | 5602 (17.4) | 164 (35.7) | <0.001 |
| **Had moderate Physical activity over past 30 days** | 22558 (43.0) | 22353 (43.2) | 205 (30.1) | <0.001 |
| **Sugar intake (g), mean (SD)** | 115.0 (69.7) | 114.7 (69.4) | 137.6 (87.1) | <0.001 |
| **Carbohydrate intake (g), mean (SD)** | 255.1 (115.5) | 254.9 (115.4) | 273.8 (124.1) | <0.001 |
| **Energy intake (KCAL), mean (SD)** | 2060.7 (883.4) | 2059.8 (883.2) | 2132.0 (890.4) | 0.060 |
| **Diabetes** | 5821 (11.1) | 5695 (11.0) | 126 (18.2) | <0.001 |
| **Cardiovascular disease <sup>b</sup>** | 4869 (9.1) | 4749 (9.0) | 120 (16.7) | <0.001 |

Note: Data are presented as frequency (%) unless specified. Numbers may not sum to totals due to missing values; percentage may not sum to 100 due to rounding. SD, standard deviation; BMI, body mass index; KCAL, kilocalories.
<sup>a</sup> Waist circumference is defined as low (men $\leq 94$ cm, women $\leq 80$ cm), high (men 94-102cm, women 80-88cm), very high (men $>102$ cm, women $>88$ cm).
<sup>b</sup> cardiovascular disease is defined as having at least one of the congestive heart failure, coronary heart disease, angina, heart attack, and stroke
P-value was for comparing characteristics between people with and without SMI.

People with SMI had a higher BMI (30.5 vs 28.6, *p*<0.001). They had a very high waist circumference (68.2% vs 52.9%, p<0.001) and were more likely to be current smokers (38.3% vs 19.4%, *p*<0.001) compared with people without SMI. More people with SMI suffered from substance misuse (35.7% vs 17.4%, p<0.001) and were less physically activity (30.1% vs 43.2%, *p*<0.001). People with SMI consumed more sugar (137.6 vs 114.7 gram, p<0.001) and carbohydrates (273.8 vs 254.9 gram, *p*<0.001), and were more likely to suffer from diabetes (18.2% vs 11.0%, *p*<0.001) and cardiovascular disease (16.7% vs 9.0%, p<0.001; Table 1)

### Oral health in people with and without SMI

For oral hygiene behaviour, there were similar rates of dental visiting between people with and without SMI, but people with SMI used dental floss less (18.6% vs 31.7% use dental floss every day, *p*<0.001). Due to the low response of tooth brushing frequency in people with SMI (*n*=3), no conclusion can be drawn for toothbrushing (Table 2).

**Table 2.**
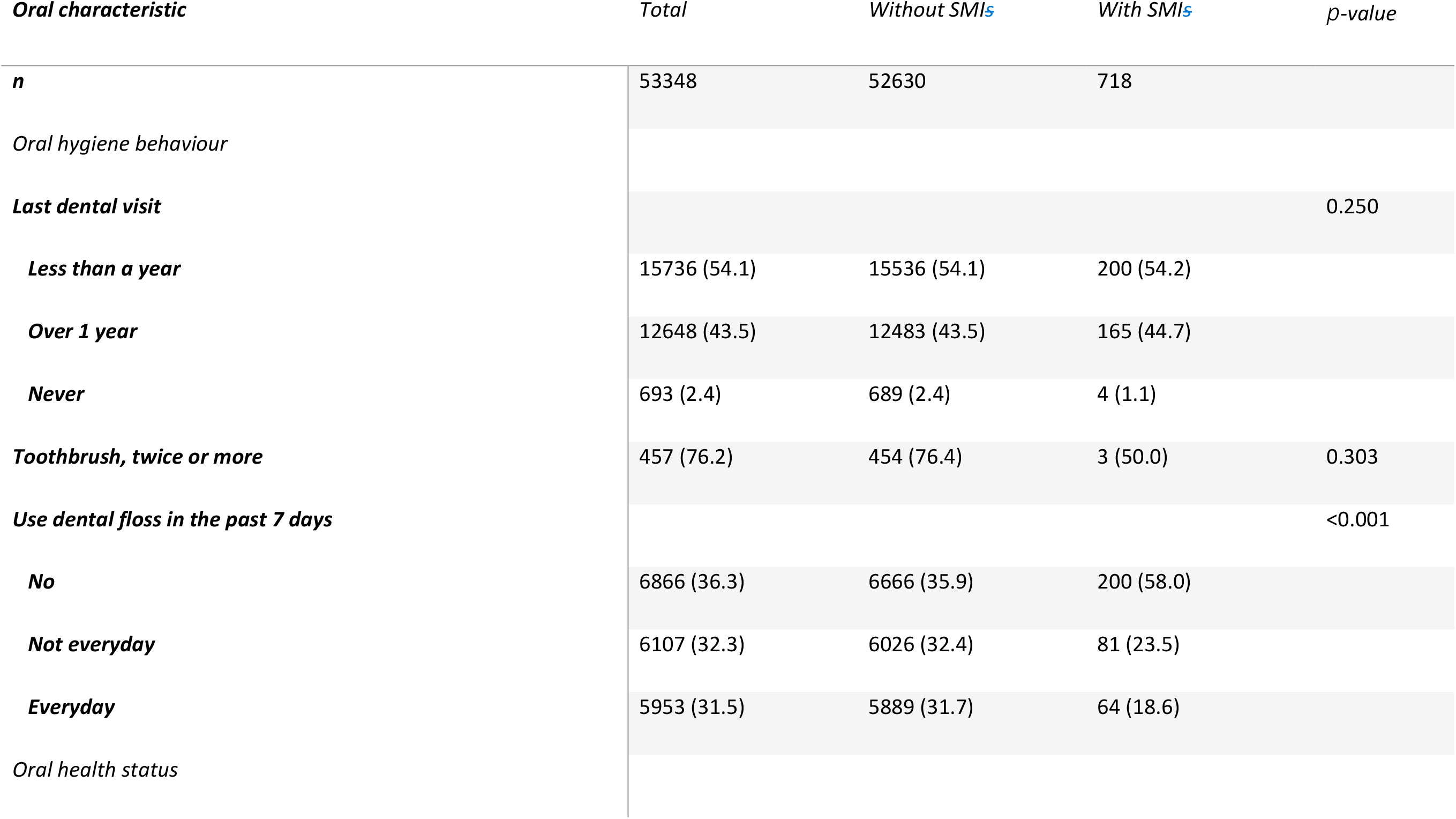
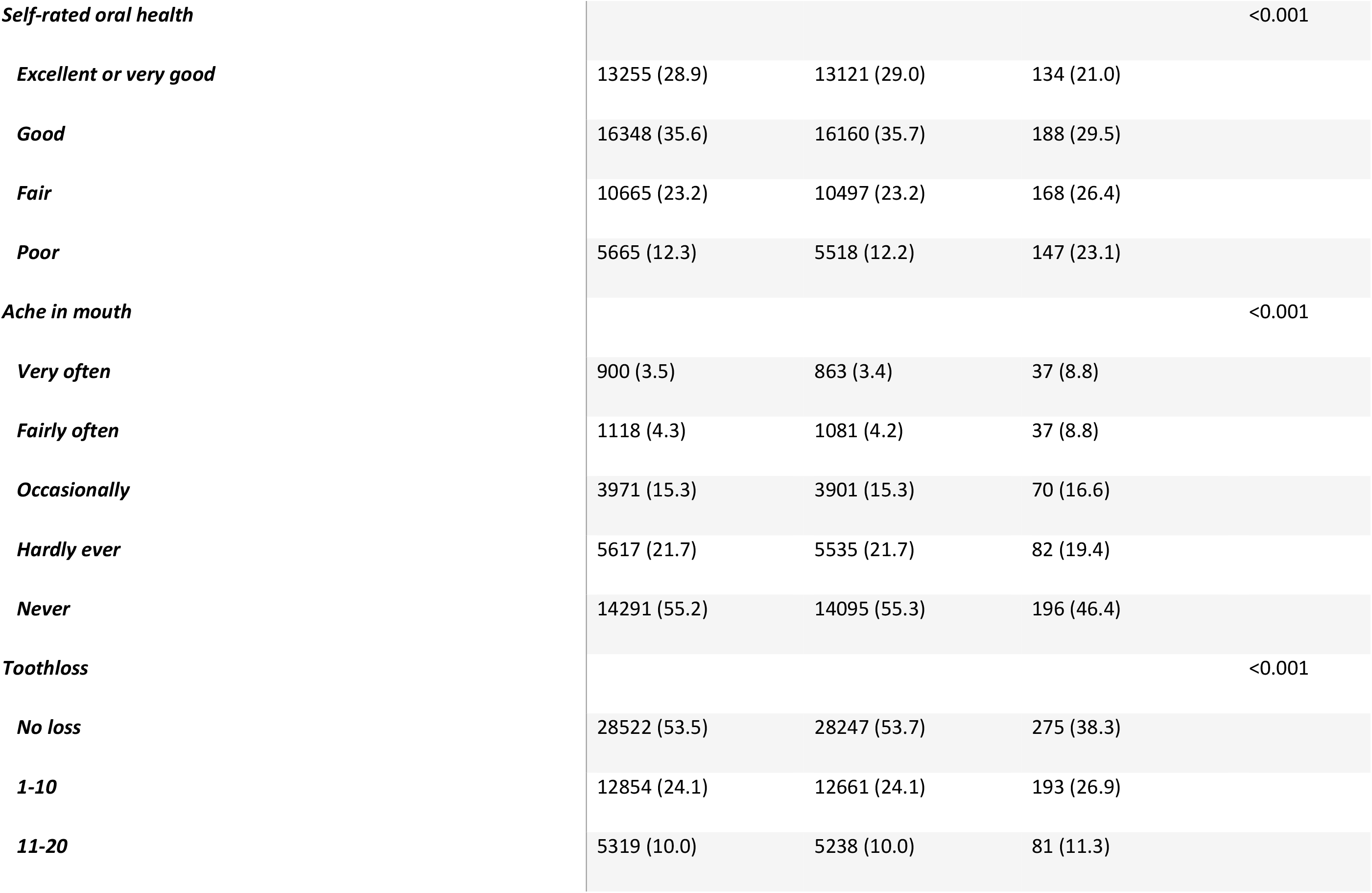
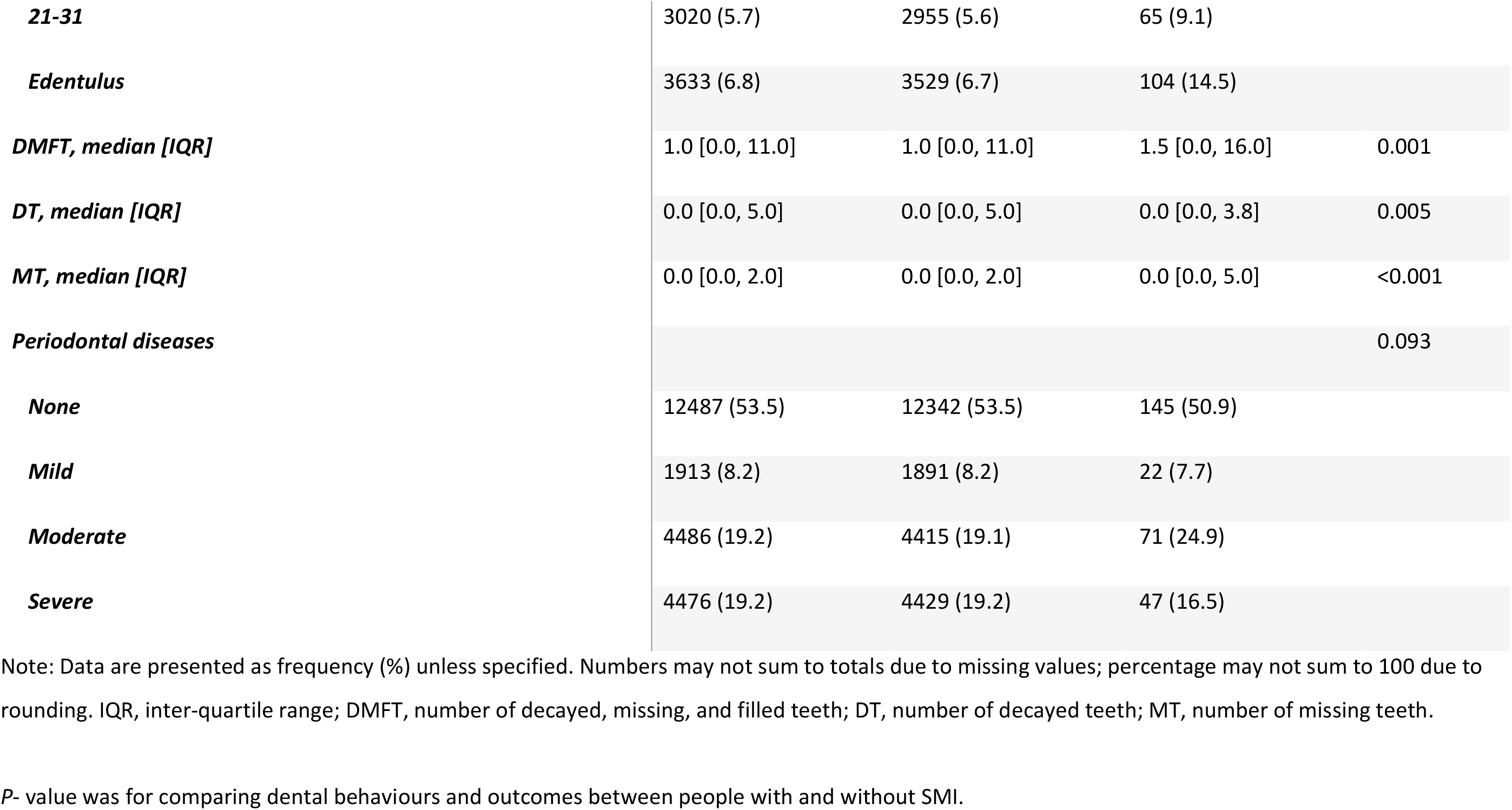
Oral hygiene behaviour and oral health status of participants with and without SMIs. NHANES (n = 53,348), 1999-2016.

For oral health outcomes, people with SMI were more likely to rate their oral health as ‘poor’ (23.1% vs 12.2%, *p*<0.001), very often had ache in mouth (8.8% vs 3.4%, *p*<0.001), being edentulous (14.5% vs 6.7%, *p*<0.001), and experienced higher level of tooth decay (DMFT median 1.5 vs 1.0, *p*=0.001). No disparity in periodontal disease levels were found. (Table 2).

Even with further adjustment of demographics lifestyles, comorbidities and oral hygiene behaviour, people with SMI were more likely to experience tooth loss (*OR* =1.40, 95% *CI* 1.12-1.75; Table 3).

**Table 3.** Association between severe mental illness status and oral health outcomes, NHANES (matched *n* = 2872), 1999-2016.

|  | Ordinal oral health outcomes (OR, 95% CI)<sup>a</sup> |  |  |  | Numeric oral health outcomes (95% CI) |  |  |  |  |  |
| --- | --- | --- | --- | --- | --- | --- | --- | --- | --- | --- |
|  | Self-rated oral health <sup>d</sup> | Ache in mouth <sup>e</sup> | Tooth loss <sup>f</sup> | Periodontal disease severity <sup>g</sup> | DMFT |  | DT |  | MT |  |
| models |  |  |  |  | Zero-inflated model, OR | Count model, IRR | Zero-inflated model, OR | Count model, IRR | Zero-inflated model, OR | Count model, IRR |
| Sample size N | 2514 | 1533 | 2872 | 1302 | 2872 |  | 2872 |  | 2872 |  |
| Unadjusted | 1.73 ***<br>(1.45-2.08) | 1.63 ***<br>(1.28-2.08) | 1.65*** (1.42-1.93) | 0.90 (0.70-1.16) | 0.94<br>( 0.79-1.11) | 1.17***<br>( 1.07-1.27) | 1.21*<br>(1.01-1.45) | 0.91<br>(0.81-1.02) | 0.81*<br>(0.65-1.00) | 1.36***<br>(1.14-1.62) |
| Adjusted for <sup>b,c</sup> |  |  |  |  |  |  |  |  |  |  |
| + demographics | 1.46 ***<br>(1.20-1.77) | 1.37 *<br>(1.06-1.78) | 1.56 ***<br>( 1.32- 1.85) | 0.86 (0.66-1.12) | 0.95 (0.79-1.13) | 1.13**<br>(1.05-1.22) | 1.18 (0.98-1.43) | 1.00<br>(0.90-1.11) | 0.85 (0.68-1.05) | 1.31***<br>(1.11-1.54) |
| + lifestyles | 1.25<br>(0.97-1.61) | 1.40 *<br>(1.01-1.93) | 1.44 **<br>( 1.16- 1.80) | 0.73 (0.53-1.01) | 0.95 (0.75-1.20) | 1.08<br>(0.97-1.19) | 1.11 (0.87-1.42) | 1.02<br>(0.90-1.17) | 0.90 (0.66-1.23) | 1.17<br>(0.94-1.46) |
| + comorbidities | 1.22<br>(0.95-1.57) | 1.38<br>(1.00-1.91) | 1.40 **<br>( 1.12- 1.75) | 0.72 (0.52-1.00) | 0.94 (0.74-1.19) | 1.06<br>(0.96-1.18) | 1.09 (0.85-1.39) | 1.04<br>(0.91-1.19) | 0.89 (0.65-1.22) | 1.13<br>(0.91-1.41) |
| + oral hygiene<br>behaviour | 1.25<br>(0.96-1.61) | 1.36<br>(0.98-1.89) | 1.40 **<br>( 1.12- 1.75) | 0.72 (0.52-1.00) | 0.94 (0.74-1.19) | 1.04<br>(0.94-1.16) | 1.09 (0.85-1.39) | 1.02<br>(0.90-1.17) | 0.85 (0.62-1.16) | 1.06<br>(0.84-1.32) |

<sup>a</sup> OR estimates and 95% CI were pooled over the 5 imputed datasets.
<sup>b</sup> logistic regression or ordinal logistic regression models were performed for ordinal or binary oral health outcomes to assess the inequality of oral health outcomes comparing people with and without SMI. Models are incrementally adjusted for demographics (age, sex, ethnicity, marital status, income), lifestyles (BMI, smoking status, substance misuse, physical activity, sugar intake), comorbidities (diabetes, cardiovascular disease), and oral health behaviour (dental visit, dental floss use).
<sup>c</sup> Zero-inflated negative binomial (ZINB) models were performed to assess the inequality of dental caries experience comparing people with and without SMI. ZINB model is a 2-part model, with zero-inflated model predict the chance of excessive zeros, and the count model predict the number of DMFT/DT/MT. For example, in DMFT unadjusted model, zero-inflated odds ratio 0.94 indicated that people with SMI has 6% (1-0.94) lower chance to be caries free (DMFT=0), and the count model IRR as 1.17 showed that people with SMI has 17% higher chance to have more dental caries compare to people without SMI.
<sup>d</sup> self-rated oral health: excellent to good (ref), fair to poor
<sup>e</sup> Ache in mouth: never or hardly ever(ref), occasionally to very often.
<sup>f</sup> tooth loss: no loss (ref), 1-10, 11-20, 21-31, and edentulous
<sup>g</sup> periodontal disease severity: none (ref), mild to moderate, severe
OR, odds ratio; IRR, incident rate ratio; DMFT, number of decayed, missing and filled teeth; DT, number of decayed teeth; MT, number of missing teeth due to decay.

### Risk factors of poor oral health in people with SMI

Finally, we explored risk factors for poor oral health outcomes in the SMI sample (Table 4). Older age (OR 1.07, 95% CI: 1.05-1.09), smoking history (OR 2.62, 95% CI 1.72-3.99), and diabetes (OR 2.05, 95% CI: 1.18-3.55) were associated higher levels of tooth loss. Conversely, higher family income (OR 0.77, 95% CI: 0.67-0.89) and using dental floss everyday (OR 0.50, 95% CI: 0.28-0.90) were associated with lower levels of tooth loss. Higher family income was found to be associated with less risk of ache in mouth (OR 0.75, 95% CI: 0.59-0.91) and less risk of poor self-rated oral health (OR 0.84, 95% CI:0.72-0.99). Engaging in physical activity halved the risk of poor self-rated oral health (OR 0.54, 95% CI 0.34-0.86). In terms of dental caries, older age, white ethnicity, being a smoker, and not using dental floss everyday were associated with a higher DMFT score. For the number of missing teeth due to decay, having full dentition (zero-inflated part of the model) was associated with younger age, white ethnicity, and higher family income. Older age, white ethnicity, smoker history, diabetes, and not using dental floss everyday were associated with a higher number of missing teeth. No risk factors were identified for periodontal disease severity potentially due to the small sample size for this outcome. The effect sizes and significance levels remained stable when various modelling strategies were applied with different covariates to test the robustness the results (Table 4).

**Table 4.**
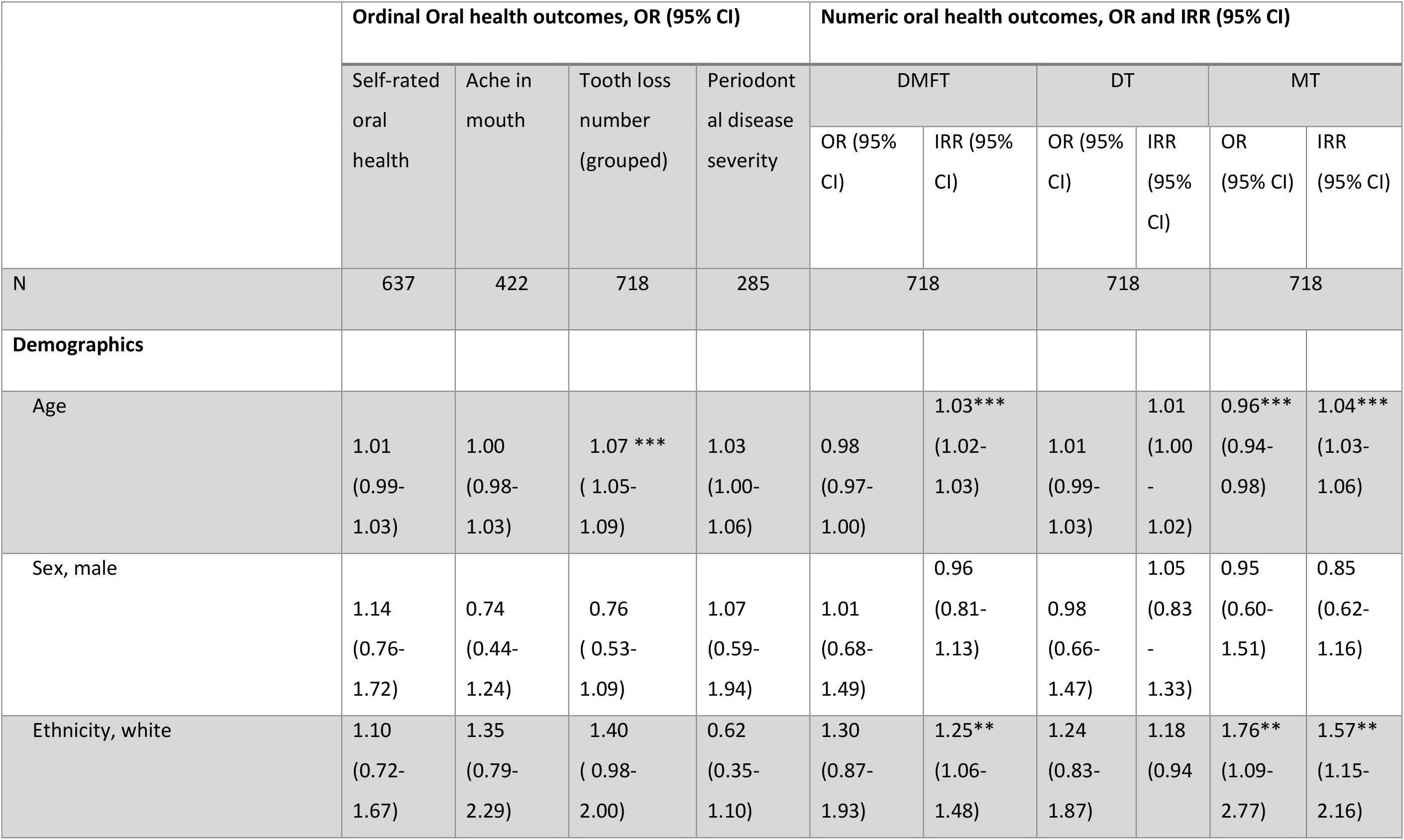
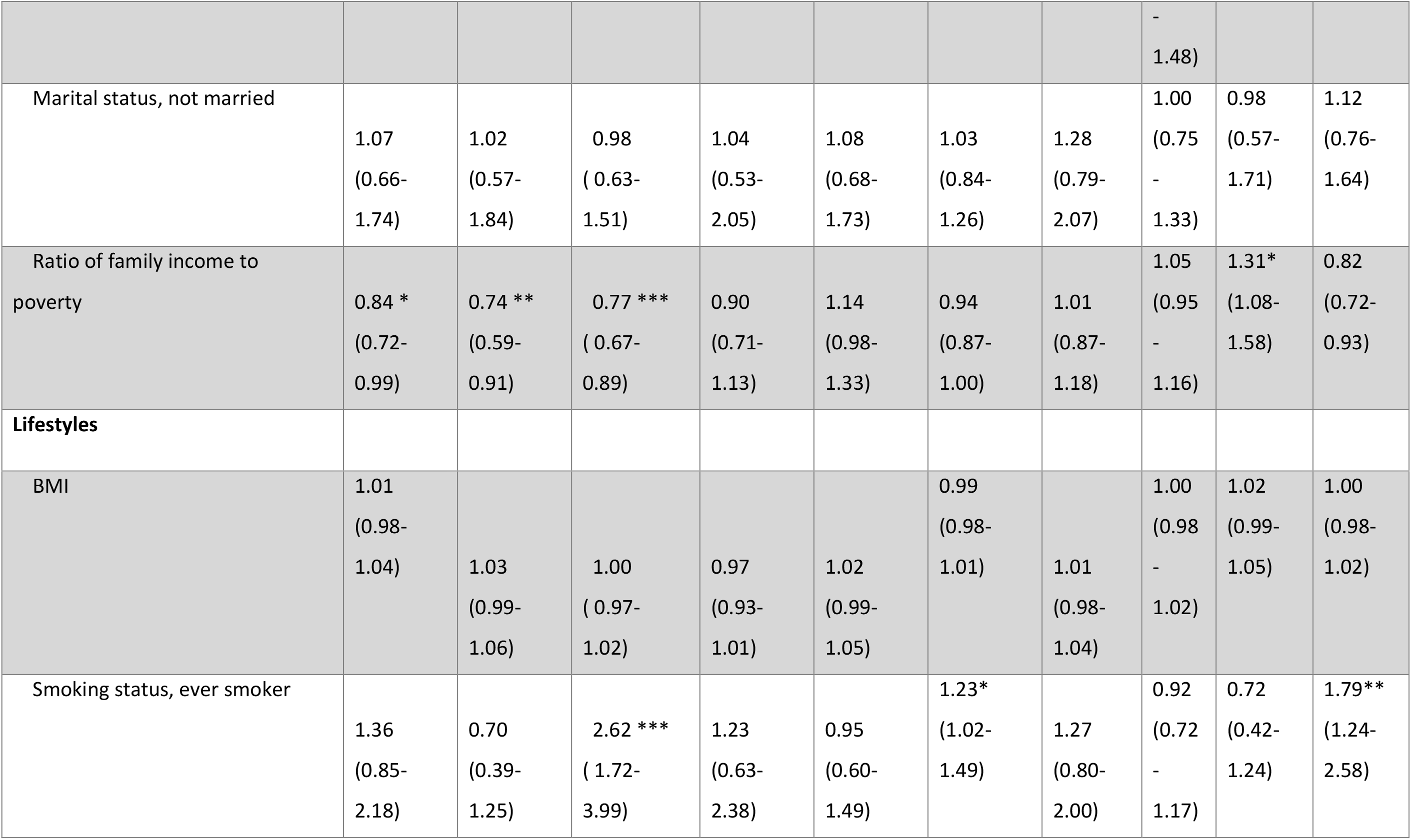
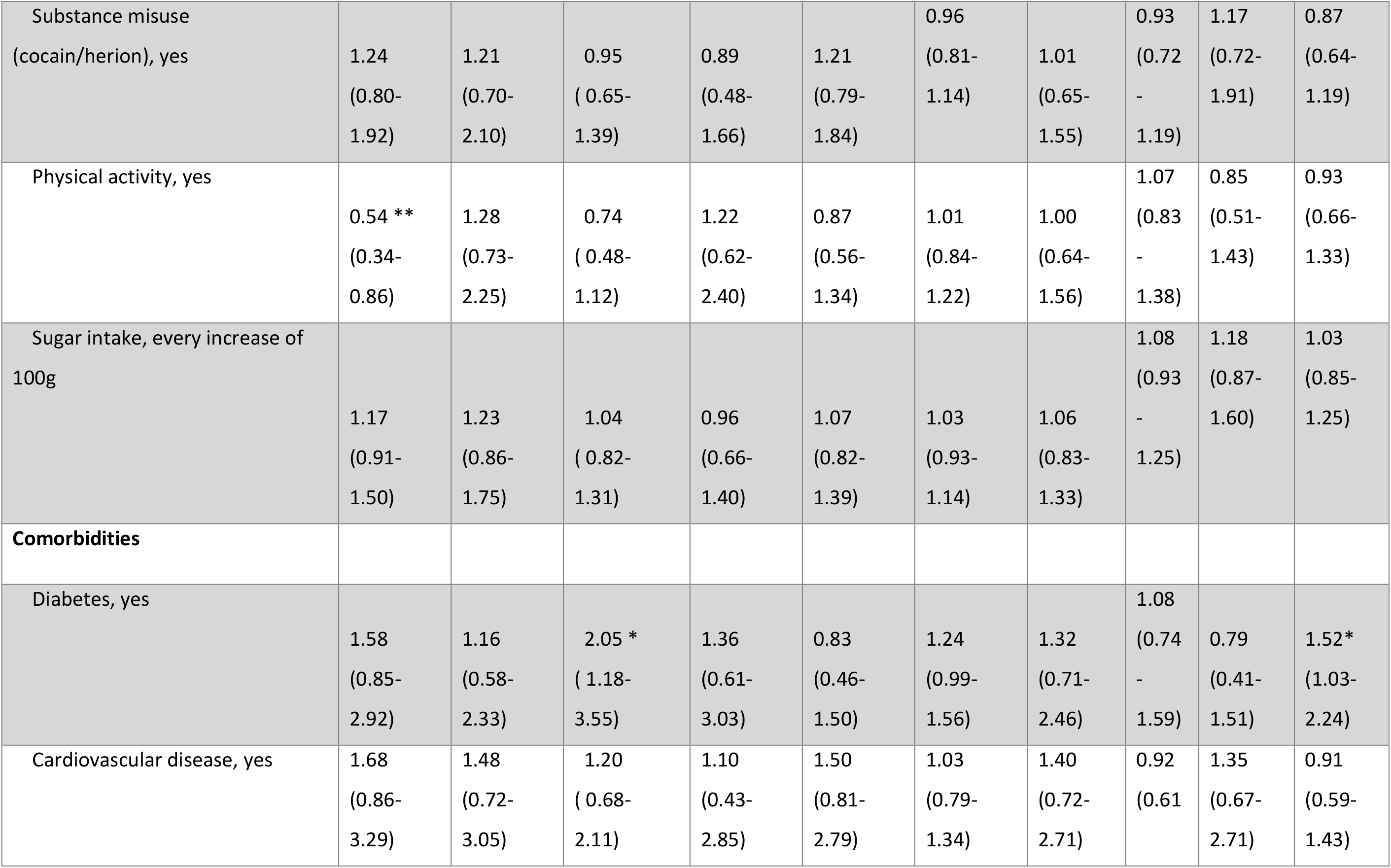
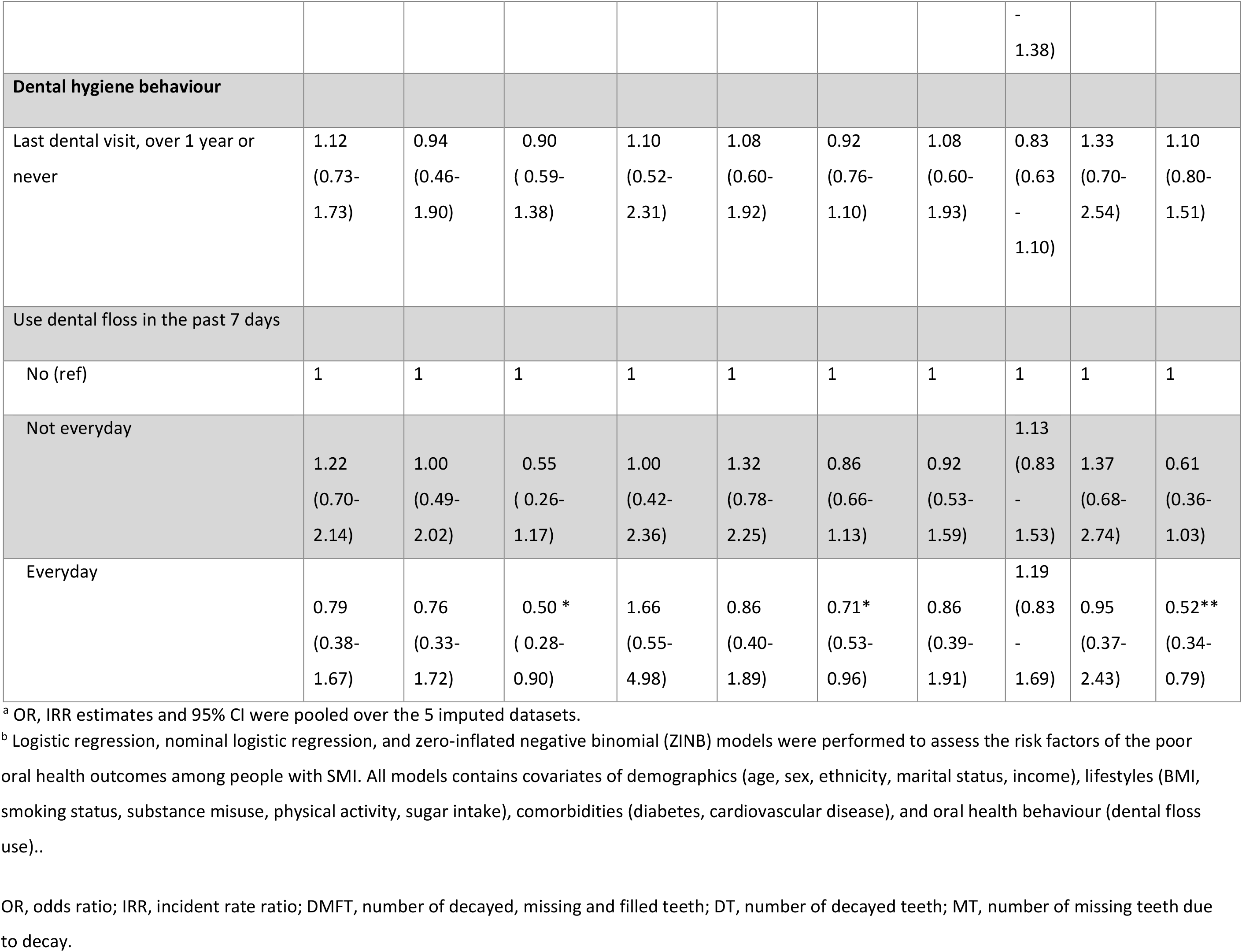
Risk factors of poor oral health in people with severe mental illness, NHANES (n = 718), 1999-2016.

## Discussion

This study aimed to investigate the inequality in oral health between people with and without SMI. We found that people with SMI suffered with more teeth lost due to dental disease, and the modifiable risk factors for their poorer oral health included smoking and poor oral hygiene habits. This epidemiological study investigated one of the largest datasets in oral health inequalities in people with SMI to date.

It was noted that disparity in oral health existed for all outcomes despite adjustments with demographics, lifestyle, comorbidities and oral hygiene behaviour, though only tooth loss showed a final significant difference between people with and without SMI potentially due to the sample size and multiple adjustment of several confounders. One should note that the effect sizes observed in each of the oral outcome were relatively large and did not show much change across the models, demonstrating the robustness of the results.

It is interesting that people with SMI were 40% more likely to loss more teeth compared to the general population and that around 15% of the sample were edentulous. Tooth loss is an end stage of periodontal disease, dental cavity, and other dental disease when preventive or conservative treatments fail (25). Tooth loss can cause considerable suffering and difficulties with essential functions such as eating and speaking (26). Additionally, the process of tooth loss can be intrusive and cause discomfort due to pain and bleeding in mouth. The long term effect of poor oral health can even lead to disfigurement, acute and chronic infection, eating and sleeping disruption leading to mental problems, finally hospitalisation or even lost work (27). People with SMI therefore require additional support around their oral health to prevent this adverse outcome from occurring.

We did not find any evidence of disparity in periodontal disease severity between people with and without SMI. This might have been due to low rates of measurement of periodontal disease in the NHANES data (44% overall), which was particularly true of people with SMI where periodontal status was only available in 283 (39%) participants. Another reason might be sampling bias in the participants—people with more SMI might be less likely to participate in those national surveys due to the difficulties in various personal reasons. We are not aware of any literature that has focused on the periodontal disease severity of people with SMI so our study is the first one examining this area. Further studies investigating the periodontal status are needed to enable us to understand the needs of people with SMI.

This study identified particular groups of people with SMI who might be of higher risk of poor oral health, including people from a white ethnicity and lower family income. This may be evidence of health intersectionality whereby certain characteristics combine to particularly disadvantage subsections of the population, which has clear relevance for oral health (28-31). Lifestyle behaviours were also found to be related to oral health. For example, smoking increased the risk of tooth loss by times and dental caries by more than 20%.(32) There is strong evidence to suggest that people with SMI are more likely to smoke and smoke heavily than the general population (12, 33), and our findings provide particular evidence of its harmful effect on oral health. Conversely, healthy lifestyle such as doing moderate physical every week and using dental floss every day was shown to reduce the chance of poor oral health by about 50%. This benefit of physical exercise and oral health has also been reported elsewhere (34), as exercise may lead to greater satisfaction with general health, mental health, which extends to oral health. Furthermore, physical exercise may protect against inflammatory process that increasingly are being recognised as mediators of physical disorders, which may also apply to oral disorders (35, 36). The benefit of flossing may offer additional protection to teeth and gums, over brushing alone (37). Important to note is that flossing in this data might also have acted as a proxy indicator of better oral hygiene behaviours more generally and greater attention to one’s teeth.

The association between tooth loss and diabetes was unsurprising given that diabetes may contribute to dental problems, especially if blood glucose control is limited, adversely affecting the oral bacterial flora and leading to tooth decay and gum disease (18, 38). This may, in part, reflect a shared exposure to common risk factors that include smoking, poor nutrition, obesity and physical inactivity. However, causality is likely to be more complex; there appears a two-way relationship in which poor glycaemic control increases the risk of periodontitis, and periodontal inflammation adversely affects glycaemic control (39). Older age was found to be associated with tooth loss and dental caries. Therefore, the early phase of mental illness may present a critical time to prevent progression to poor oral health we observed in older age.

### Limitations and future research

This study used a proxy measure of SMI in prescriptions of antipsychotic or mood stabilising medication. Past studies have indicated that they accurately map onto diagnosis of SMI (40). However, the overlap between medication and formal diagnostic constructs in the current dataset are unclear. Linked medical record and clinical diagnosis would offer more accurate identification of people with SMI, and future research using such data will offer stronger evidence in this area. Due to small numbers and missing data in some of the years of NHANES data collection, we were unable to investigate the impact of polypharmacy and Clozapine use, which may have side effects that adversely affect oral health. Psychological factors (e.g. dental anxiety) and systemic risk factors (e.g. access to specialist services) also require further investigation in relation to poor oral health in SMI.

### Clinical implications

Our findings highlight the importance of tackling oral health inequalities in people with SMI. In the United Kingdom, the recent National Institute of Clinical Excellence Rehabilitation Guidelines for complex psychosis (41) recommend that mental health services are aware of and signpost to support around oral health, but this appears to be the exception rather than the rule. Our research suggests that oral health inequalities extend to community samples, which also needs recognition in broader mental health and dental policy and guidelines.

Interventions around oral health in SMI have largely focused on education. The Three Shires Trial (42) evaluated brief dental awareness training for mental health staff working in early intervention for psychosis services, which had no significant impact on any outcome. Other small feasibility studies have suggested that education interventions may oral health outcomes in SMI (e.g. plaque) when combined with behavioural change techniques (43). Little is known about the effectiveness of such interventions when rolled out in routine clinical practice, and further research is required in this area.

Our findings help to inform an ‘at risk profile’ for poor oral health in people with SMI. For example, people with diabetes and smokers may be at particular risk and therefore benefit from additional support and signposting around their oral health. There exist focused evidence-based smoking cessation programmes for people with SMI (33), which also require greater uptake across healthcare providers. Lastly, the effect of age on tooth loss, the number of missing teeth and DMFT score may suggest opportunities for early detection and intervention around oral health for younger people in an early phase of psychosis.

## Conclusion

To summarise, we found that people with SMI are more likely to experience toothloss than people without SMI in the general population. Risk factors for poor oral health in this population included older age, white ethnicity, lower income, smoking history, and diabetes. Physical activity and daily use of dental floss were associated with better oral health outcomes. Overall, the findings highlight oral health inequalities in people with SMI, which require further attention in clinical settings and future research.

## Data Availability

The NHANES 1999-2016 data is available at CDC website: https://www.cdc.gov/nchs/nhanes/index.htm, and is accessible and free to download for everyone.

## Funding

This project has been funded by the Closing the Gap network. Closing the Gap is funded by UK Research and Innovation and their support is gratefully acknowledged (Grant reference: ES/S004459/1). Any views expressed here are those of the project investigators and do not necessarily represent the views of the Closing the Gap network or UKRI.

## Conflict of Interest Statement

The authors declare no conflict of interest.

## Conflict of interest

The authors declare no conflicts of interest with respect to the authorship and/or publication of this article.

## Appendix I

a. Drug code used to identify SMI patients as long as at least 1 of the following medication is taken:

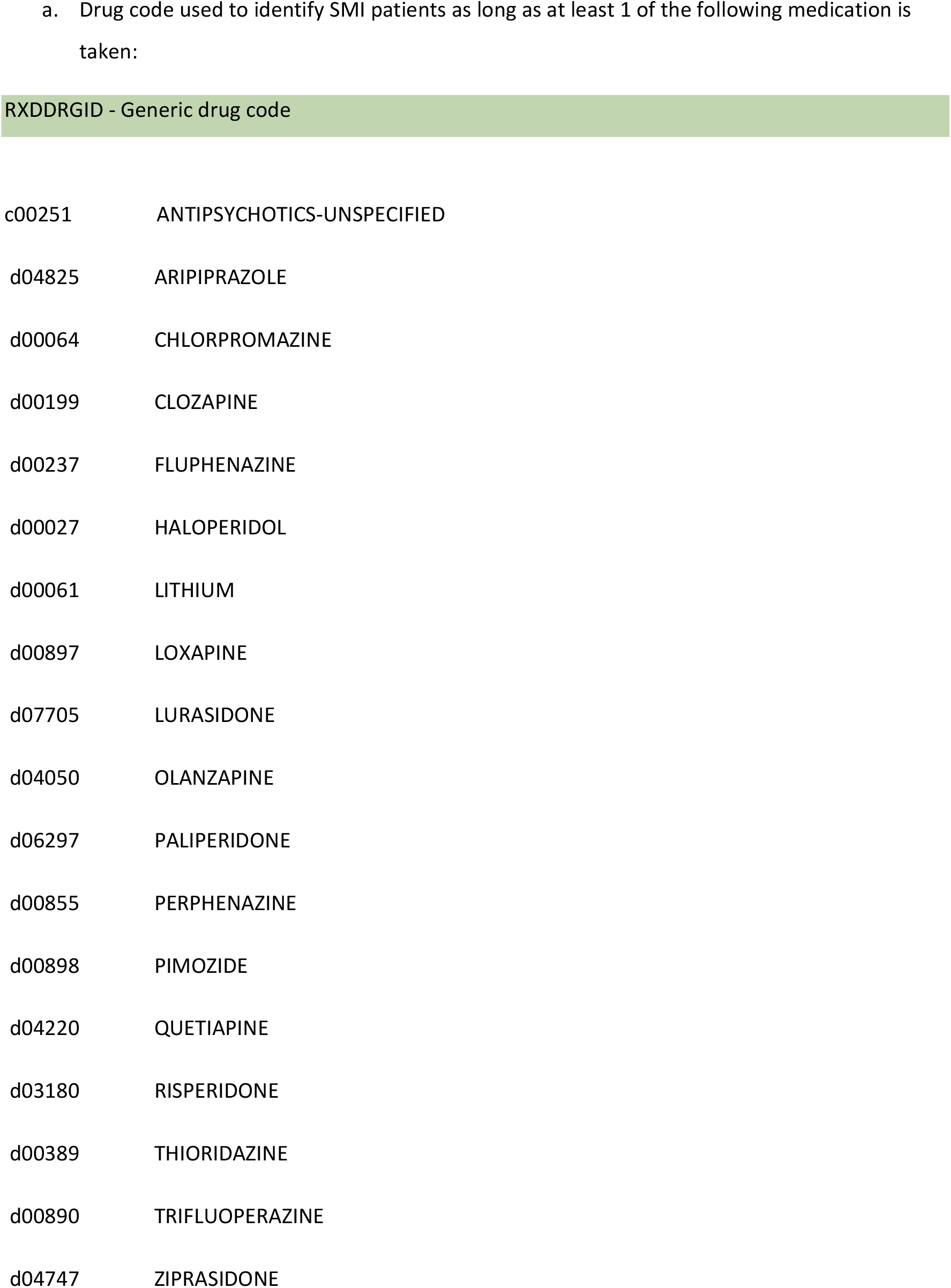

b. Drug code validation using ICD-10-CM code: “RXDRSC1” is the ICD10CM code, use F31.9(bipolar), F29 (psychosis), F20(schizophrenia)

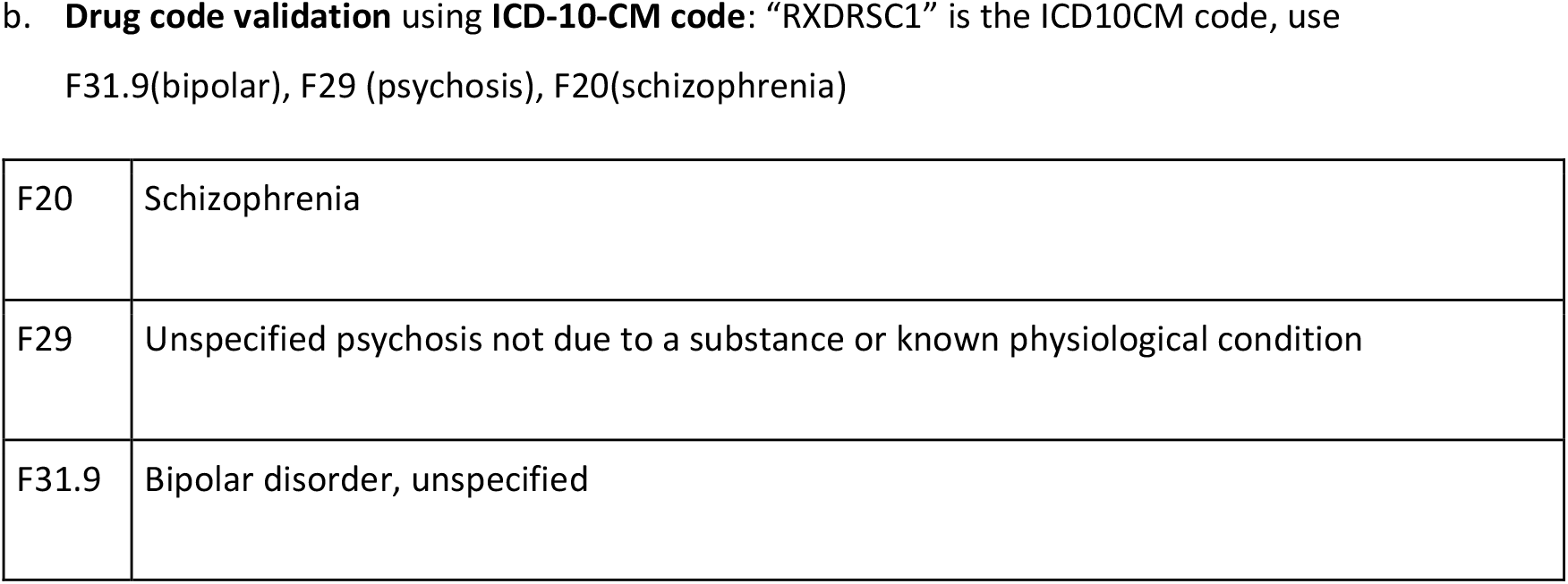

## Notes

### Competing Interest Statement

The authors have declared no competing interest.

